# Prospective Characterization of Progressive Supranuclear Palsy in a Multi-Ethnic Asian Cohort

**DOI:** 10.1101/2022.10.01.22280553

**Authors:** Shen-Yang Lim, Alfand Marl F. Dy Closas, Ai Huey Tan, Jia Lun Lim, Yi Jayne Tan, Yuganthini Vijayanathan, Yi Wen Tay, Raihanah binti Abdul Khalid, Wai Keong Ng, Ruban Kanesalingam, Pablo Martinez-Martin, Jia Nee Foo, Weng Khong Lim, Adeline Su Lyn Ng, Eng-King Tan

## Abstract

**Background:** Progressive supranuclear palsy (PSP) is a rare, disabling, neurodegenerative disease. There are limited studies on the spectrum of PSP predominance-types and their clinico-demographic features among Asian patients. We prospectively characterized the clinical features, disease severity, and caregiver burden in a multi-ethnic Asian PSP cohort.

**Methods:** Consecutively-recruited patients with PSP (n=104, 64.4% male; 67.3% Chinese, 21.2% Indians, 9.6% Malays) were extensively phenotyped by a movement disorders neurologist using the MDS-PSP clinical diagnostic criteria and PSP-Clinical Deficits Scale (PSP-CDS). Caregiver burden was measured using the modified Zarit Burden Interview (ZBI). Investigations were reviewed to help rule out potential PSP mimics.

**Results:** There were 104 patients, consisting of 48.1% Richardson syndrome (PSP-RS), 37.5% parkinsonian phenotype (PSP-P), and 10.6% progressive gait freezing phenotype (PSP-PGF). Mean age at motor onset was 66.3±7.7 years, with no significant differences between the PSP phenotypes. Interestingly, REM-sleep behaviour disorder (RBD) symptoms and visual hallucinations (considered rare in PSP) were reported in 23.5% and 22.8% of patients, respectively, and a family history of possible neurodegenerative or movement disorder in 20.4%. PSP-CDS scores were highest (worst) in PSP-RS; and correlated moderately with disease duration (r_s_=0.45, P<0.001) and weakly with caregiver burden (r_s_=0.22, P=0.029) in the overall cohort. Three of 48 (6.3%) patients who had whole-exome sequencing harboured pathogenic/likely pathogenic *GBA* variants.

**Conclusions:** This prospective characterization of a relatively large cohort of Asian PSP patients depicts significant heterogeneity in clinical features and disease burden. Unexpectedly high rates of RBD symptoms, visual hallucinations, and familial involvement warrant further study.

## 1. Introduction

Progressive supranuclear palsy (PSP) is a debilitating neurodegenerative disorder affecting ≈5-10 people per 100,000 population [1]. In clinical practice, it is probably the most common of the “Parkinson-plus” syndromes, and variants other than the classical Richardson syndrome (PSP-RS), such as PSP with predominant parkinsonism (PSP-P) and PSP with predominant progressive gait freezing (PSP-PGF), are also increasingly recognized [1,2]. The clinical diagnoses of the various PSP phenotypes were formalized by the International Parkinson and Movement Disorder Society (MDS)-endorsed PSP study group in 2017 [3], anticipating that a better understanding of PSP variants will improve patient management (e.g., in terms of prognostication) and research (e.g., facilitating earlier diagnosis, and recruitment into studies). However, to our knowledge, there has been no published research on the prospective characterization of Asian PSP patients using the MDS-PSP diagnostic criteria.

Another unmet need in the field, until very recently, has been the lack of a simple rating scale to grade PSP severity that can be easily applied in routine clinical settings [4]. In 2020, the MDS-endorsed PSP study group published on a brief rating scale that captures many of the pertinent clinical aspects of the disease with relevance to activities of daily living [4]. This instrument could be administered by the original group of experts who developed it in 4 minutes on average. It was also found to be clinimetrically sound for clinical care and research, and has been proposed to be equally applicable to PSP-RS as well as to variant PSP phenotypes [4]. There is, however, no published literature yet on the utilization of this scale by independent researchers.

Our primary objective was to characterize PSP cases in a multi-ethnic Asian cohort by careful clinical assessments, including the application of the MDS-PSP clinical diagnostic criteria and the PSP-CDS in a routine clinic practice setting.

## 2. Subjects and methods

### 2.1 Subject recruitment

Patients and caregivers were consecutively recruited as part of an ongoing study investigating genotype-phenotype correlations in movement disorders at the University of Malaya Medical Centre, Kuala Lumpur, Malaysia. The study has been approved by the institutional ethics committee and all patients provided written informed consent. The recruitment period was from May 2020 to August 2022.

### 2.2 Clinical assessments

All patients were clinically assessed by an experienced movement disorder neurologist (SYL) during routine outpatient clinic consultations. Standard demographic and clinical data were collected, including information on: current age; ancestry/ethnicity; disease duration from motor symptom onset as well as from the time of diagnosis of a movement disorder; possible history of neurodegenerative disease or movement disorder (e.g., Parkinson’s disease [PD], early-onset dementia (<65 years), or motor neuron disease) in the immediate or extended family; and medication treatment including levodopa equivalent daily dose (LEDD) and use of dopamine agonists and amantadine. Patients and caregivers were also systematically queried about the presence of a history of REM-sleep behaviour disorder (RBD) symptoms and visual hallucinations; where possible, video-documentation of their testimony was recorded.

Using the 2017 MDS-PSP clinical diagnostic criteria, patients were categorized into various predominance-types of PSP, with differing levels of diagnostic certainty (“probable”, “possible” or “suggestive of”) [3]. The medical charts for all patients were reviewed and their PSP categorization rechecked by a movement disorders fellow (AMD); any disagreements with the original designations were resolved by discussion with SYL. Patient clinical details were kept confidential and patient ID were not made known to others outside of the research group. The PSP-CDS evaluates seven clinical domains (“Akinesia-rigidity”, “Bradyphrenia”, “Communication”, “Dysphagia”, “Eye movements”, “Finger dexterity”, and “Gait and balance”) [4]. Each item is scored from 0-3 (0=No deficit; 1=Mild; 2=Moderate; 3=Severe), with descriptions provided for each category in the scoring table (Figure 1 in the original publication); further detailed explanations were also given in Supplement 1 accompanying the publication [4]. Therefore, higher scores indicate greater disease severity.

A modified ZBI, widely used to assess caregiver burden in PD [5,6], was given to the patient’s primary caregiver to self-complete after the consultation. English or Chinese versions were available, according to the caregiver’s preference. This instrument consists of 24 items assessing the caregiver’s feelings, each item scoring from 0 (Never) to 4 (Nearly always). Thus, higher scores indicate greater burden.

### 2.3 Review of investigations

Brain scans were assessed by SYL and AMD to rule out PSP mimics [3,7], and to qualitatively determine the presence of the “hummingbird sign” on mid-sagittal T1-weighted image [1]. Additionally, the results of whole-exome sequencing (WES) as well as testing for repeat expansion in *C9orf72*, done for a subset of patients in this report as part of another study in collaboration with the National Neuroscience Institute, Singapore, were reviewed to help rule out conditions potentially masquerading as PSP (e.g., Parkinson’s disease [PD], dementia with Lewy bodies [DLB], frontotemporal dementia, Alzheimer’s disease, and inherited vasculopathies) [3,7].

### 2.4 Statistical analysis

Descriptive statistical analysis was performed for the demographic and clinical features of the entire cohort of PSP patients and the three most common forms (PSP-RS, PSP-P and PSP-PGF). Normality tests were conducted using the Shapiro-Wilk test. Between-group comparisons were conducted using Chi-square test (for categorical data), one-way ANOVA (for normally distributed data), and Kruskal-Wallis test (for non-normally distributed data). Spearman correlations were analyzed between scores of the PSP-CDS vs. the ZBI and clinico-demographic parameters including age, disease duration, and LEDD. Correlations were considered weak for r_s_=0.10-0.29 and moderate when r_s_=0.30-0.49. *Post hoc* analyses (e.g., comparing age, disease duration, LEDD, use of amantadine or dopamine agonist, and ZBI scores between those with, vs. those without, visual hallucinations) were done using unpaired *t*-test, Mann-Whitney U test, and Chi-square test. Statistical significance was set at P<0.05.

## 3. Results

### 3.1 Subject recruitment

All 104 patients participating in the study were classified using the MDS-PSP diagnostic criteria. All had PSP-CDS ratings completed during the clinic consultation, and took around 5-10 minutes. Evaluations were done in-person in the outpatient clinic, except for two patients who were severely disabled and could not travel interstate/overseas during the COVID19 pandemic. These patients with long-standing probable PSP-RS had been seen multiple times in the past, in person, and could be adequately assessed via video-conferencing. ZBI forms were completed by 99 caregivers (not returned in four cases; and one patient who was still very independent in his activities of daily living did not have a caregiver).

### 3.2 Frequency of PSP predominance-types

The most frequently encountered PSP predominance-types were PSP-RS (48.1% of cases), followed by PSP-P (37.5%) and PSP-PGF (10.6%) (Table 1). There was only one case for each of the following PSP predominance-types: Possible PSP-CBS (corticobasal syndrome), Suggestive of PSP-PI (postural instability); Suggestive of PSP-SL (speech-language predominance); and PSP-Cerebellar type (PSP-C).

**Table 1.** Clinico-demographic characteristics, disease severity, and caregiver burden in the overall PSP cohort and the three main PSP subgroups

|  |  | Overall PSP cohort | PSP-RS | PSP-P | PSP-PGF | P value |
| --- | --- | --- | --- | --- | --- | --- |
| **Number of Patients** |  | 104 | 50 (48.1%) | 39 (37.5%) | 11 (10.6%) | - |
| *Degree of diagnostic certainty* |  |  |  |  |  |  |
| Probable |  |  | 50 | 31 | 4 |  |
| Possible |  |  | 0 | N/A | 7 |  |
| Suggestive of |  |  | 0 | 8 | N/A |  |
| **Age (years)** |  | 71.8 ± 7.4 | 70.6 ± 7.0 | 72.2 ± 7.5 | 75.3 ± 6.9 | 0.134<sup>a</sup> |
| **Gender (%male)** |  | 67 (64.4%) | 30 (60.0%) | 28 (71.8%) | 6 (54.5%) | 0.406<sup>b</sup> |
| **Ethnicity** | % Chinese | 70 (67.3%) | 33 (66.0%) | 27 (69.2%) | 7 (63.6%) | 0.500<sup>b</sup> |
|  | % Malay | 10 (9.6%) | 4 (8.0%) | 4 (10.3%) | 2 (18.2%) |  |
|  | % Indian | 22 (21.2%) | 13 (26.0%) | 6 (15.8%) | 2 (18.2%) |  |
|  | % Others | 2 (1.9%) | 0 (0.0%) | 2 (5.1%) | 0 (0.0%) |  |
| **Age at motor onset (years)** |  | 66.3 ± 7.7 | 66.4 ± 7.0 | 65.1 ± 8.6 | 68.8 ± 6.7 | 0.360<sup>a</sup> |
| **Age at diagnosis (years)<sup>#</sup>** |  | 67.7 ± 8.0 | 67.8 ± 7.2 | 66.4 ± 9.0 | 69.9 ± 6.9 | 0.398<sup>a</sup> |
| **Disease duration from diagnosis (years)** |  | 3.3 [4.3] | 2.5 [2.4] | 4.9 [7.1] | 5.2 [5.6] | **0.004**<sup>*,c</sup> |
| **Disease duration from motor onset (years)** |  | 4.7 [4.3] | 4.0 [2.0] | 6.6 [6.1] | 6.2 [5.7] | **0.002**<sup>*,c</sup> |
| **% with family history<sup>##</sup> (n=103)** |  | 21 (20.4%) | 11 (22.0%) | 8 (21.1%) | 2 (18.2%) | 0.961<sup>b</sup> |
| **% on oral dopamine replacement therapy** |  | 85 (81.7%) | 38 (77.6%) | 34 (89.5%) | 9 (81.8%) | 0.341<sup>b</sup> |
| **LEDD (mg/day)** |  | 400.0 [400.0] | 393.8 [481.5] | 400.0 [300.0] | 487.5 [225.0] | 0.160<sup>c</sup> |
| **% on amantadine** |  | 31 (29.8%) | 14 (28.0%) | 11 (28.2%) | 4 (36.4%) | 0.850<sup>b</sup> |
| **Amantadine daily dose (mg/day)** |  | 0 [138] | 0 [113] | 0 [100] | 0 [200] | 0.862<sup>c</sup> |
| **% with REM sleep behaviour disorder symptoms (n=102)** |  | 24 (23.5%) | 14 (28.6%) | 9 (23.7%) | 1 (9.1%) | 0.394<sup>b</sup> |
| **% with visual hallucinations (n=101)** |  | 23 (22.8%) | 9 (18.0%) | 11 (28.9%) | 2 (22.2%) | 0.478<sup>b</sup> |
| **PSP-CDS (range of scale 0-21)** |  | 10.0 [7.0] | 11.0 [7.0] | 10.0 [6.0] | 6.0 [6.0] | **0.004**<sup>*,c</sup> |
| **ZBI (range of scale 0-96) (n=99)** |  | 33.0 ± 15.7 | 36.7 ± 14.5 | 28.4 ± 16.6 | 29.2 ± 15.0 | **0.041**<sup>*,a</sup> |

<sup>#</sup>Denotes the age at the time of diagnosis of a movement disorder. <sup>##</sup>Denotes a possible history of neurodegenerative or movement disorder in the immediate or extended family. Unless otherwise stated, each parameter in column 1 was analyzed based on the full dataset (n=104). Normally-distributed data are expressed as mean±standard deviation, non-normally distributed data as median [inter-quartile range]. <sup>a</sup>Analyzed using one-way ANOVA; <sup>b</sup>Analyzed using Chi-square; <sup>c</sup>Analyzed using Kruskal-Wallis Test. \*Denotes statistical significance. LEDD=Levodopa equivalent daily dose; N/A=Not applicable; PSP-CDS=Progressive Supranuclear Palsy Clinical Deficits Scale; REM=Rapid eye movement; ZBI=Zarit Burden Interview.

### 3.3 Clinico-demographic features

These are tabulated in Table 1 for the overall cohort, and for the three most common predominance-types (PSP-RS, PSP-P, and PSP-PGF). Individual-level data are presented in Supplementary Table 1. With only one exception, all patients were seen with an informant(s) (family member and/or caregiver), who helped to corroborate the history. There was a significant between-group difference for disease duration, which was shortest for PSP-RS. RBD symptoms and visual hallucinations were endorsed in 24/102 (23.5%) and 23/101 (22.8%) of cases, respectively (12/100=12.0% with both). However, pharmacological treatment was administered in only one and five cases, respectively, consisting of melatonin and quetiapine (25mg/d in 3; 50mg/d in 2; none warranted clozapine treatment or inpatient psychiatric management). Video-recordings of patient and/or caregiver testimonies and depictions of these phenomena, which have been considered rare in PSP [3].

A family history of possible neurodegenerative or movement disorder was recorded in 21/103 (20.4%) of cases, consisting of: 16 patients with family member(s) having PD or query PD; one having a brother with “shaking and falls”; one having a cousin with action tremor; and two having a family member with motor neuron disease. The mean age of disease onset in patients with, vs. those without, positive family history was not different (67.6±6.7 vs. 66.0±8.0 years, P=NS).

### 3.4 Rating scales: Progressive Supranuclear Palsy Clinical Deficits Scale and Zarit Burden Interview

Median PSP-CDS score for the overall cohort was 10.0 (inter-quartile range [IQR] 7.0), and ranged between 3 and 21. PSP-CDS scores were significantly different between the predominance-types (highest/worst in PSP-RS, with *post hoc* analyses showing significantly higher scores in PSP-RS vs. PSP-PGF) (Table 1). Mean ZBI score for the overall cohort was 33.0 ± 15.7, and ranged between 4 and 75. ZBI scores were also significantly different between groups (highest/worst in PSP-RS, with *post hoc* analyses showing significantly higher scores in PSP-RS vs. PSP-P).

### 3.5 Correlations and associations

A moderate positive correlation was found between scores of the PSP-CDS and disease duration (r_s_=0.45, P<0.001), whereas the correlation was weak with LEDD (r_s_=0.22, P=0.026) and the ZBI (r_s_=0.22, P=0.029) (Table 2).

**Table 2.** Correlations between Progressive Supranuclear Palsy Clinical Deficits Scale (PSP-CDS) scores, Zarit Burden Interview (ZBI) scores, and clinico-demographic characteristics of patients

|  | PSP-CDS |  | ZBI |  | LEDD |  |
| --- | --- | --- | --- | --- | --- | --- |
|  | **r<sub>s</sub>** | **P value** | **r<sub>s</sub>** | **P value** | **r<sub>s</sub>** | **P value** |
| PSP-CDS | - | - | **0.22** | **0.029\*** | **0.22** | **0.026\*** |
| ZBI | **0.22** | **0.029\*** | - | - | -<br>0.07 | 0.470 |
| Age | 0.08 | 0.441 | -0.16 | 0.107 | 0.14 | 0.155 |
| Age at motor onset | -<br>0.11 | 0.257 | -0.16 | 0.122 | -<br>0.04 | 0.715 |
| Disease duration (from motor onset) | **0.45** | **<0.001\*** | 0.05 | 0.654 | **0.39** | **<0.001\*** |
| LEDD | **0.22** | **0.026\*** | -0.07 | 0.470 | - | - |

Data were analyzed using Spearman correlations. LEDD=Levodopa equivalent daily dose; PSP-CDS=Progressive Supranuclear Palsy Clinical Deficits Scale; ZBI=Zarit Burden Interview. \*Denotes statistical significance.

The presence of a history of visual hallucinations correlated moderately with a history of RBD symptoms, with r_s_=0.38 (P<0.001). Twelve of 23 (52.2%) patients with hallucinations had RBD, and 12/24 (50.0%) patients with RBD reported hallucinations. Age (71.4±8.4 years vs. 71.8±7.1), disease duration (4.9 [5.2] vs. 4.7 [4.9]), amantadine use (13.0% vs. 33.3%), dopamine agonist use (8.7% vs. 12.8%), LEDD (450mg/d [200.0] vs. 400.0 [400.0]), PSP-CDS (9.0 [6.0] vs. 10.0 [7.0]), and ZBI (37.6±16.2 vs. 31.1±15.4) were not significantly different in patients with, vs. those without, a history of hallucinations. Most of the patients (18/23=78.3%) with a history of visual hallucinations were taking neither amantadine nor dopamine agonist.

### 3.6 Investigation findings

Brain MRI scans were available for our review in 89 patients (85.6%) and brain CT scan in another eleven (10.6%). Mid-sagittal T1-weighted image was available for review in 83 (79.8%) of cases, of whom 33 (39.8%) demonstrated a “hummingbird” sign (66.3% if cases with flattening of the superior midbrain border were also included) [1]. Brain MRI findings that were considered unusual or unexpected for PSP (structural or vascular lesions, asymmetric cerebral atrophy, or disproportionate ventriculomegaly) were observed in nine patients (Table 3) (motor symptom onset ranging 50-81 years; 3 probable PSP-RS, 3 probable PSP-P, 1 possible PSP-PGF, 1 suggestive of PSP-P, and 1 suggestive of PSP-SL). Two cases initially classified clinically as possible PSP-RS had prominent neuroimaging findings suggestive of cerebrovascular disease (Table 3) and were excluded from this cohort.

**Table 3.** Cases with unusual or unexpected brain MRI findings and/or mutations in neurodegeneration-related genes. Except for the last two patients who were excluded from the cohort (bottom rows), overall features were considered to be still in keeping with a diagnosis of progressive supranuclear palsy.

| Patient # | Unusual brain MRI findings | Relevant WES findings* | Family history | PSP classification; OPAC** core clinical features | RBD symptoms | Visual hallucinations |
| --- | --- | --- | --- | --- | --- | --- |
| 1. Patient #76 | ✓<br>Ponto-cerebellar atrophy | N/A | ✗ | Prob. PSP-RS<br>O1, P1, A2, C0 | ✓ | ✗ |
| 2. Patient #11 | ✓<br>Ventriculomegaly (but clinically unimproved after VP shunt) | N/A | ✓<br>(relative reported to have late-onset PD) | S.o. PSP-P<br>O0, P0, A2, C0 | ✓ | ✗ |
| 3. Patient #56 | ✓<br>Moderate-to-severe fronto-temporo-parietal atrophy, worse on right | ✓<br>*GBA* p.V499M LP | ✗ | Prob. PSP-RS<br>O1, P1, A2, C1 | ✗ | ✗ |
| 4. Patient #28 | ✓<br>Moderate fronto-parietal atrophy, worse on left | N/A | ✗ | S.o. PSP-SL<br>O0, P1, A2, C1 | ✗ | ✗ |
| 5. Patient #16 | ✓<br>Moderate parietal atrophy, worse on right; bilateral putaminal T2 hypointensity | ✗ | ✗ | Poss. PSP-PGF<br>O0, P0, A2, C0 | ✗ | N/A |
| 6. Patient #65 | ✓<br>Moderate parietal atrophy, worse on left | ✗ | ✓<br>(relative with MND) | Prob. PSP-P<br>O2, P0, A3, C0 | ✗ | ✗ |
| 7. Patient #52 | ✓ | ✗ | ✗ | Prob. PSP-P | ✗ | ✗ |
|  | Encephalomalacia in left frontal subcortical region |  |  | O1, P0, A1, C0 |  |  |
| 8. Patient #72 | ✓<br>Large biparietal arachnoid cysts, worse on left | N/A | ✗ | Prob. PSP-RS<br>O2, P1, A2, C3 | ✗ | ✗ |
| 9. Patient #89 | ✓<br>Moderate, confluent periventricular white matter T2 hyperintensity (note: had history of HTN) | N/A | ✗ | Prob. PSP-P<br>O2, P0, A2, C0 | ✗ | ✗ |
| 10. Patient #18 | ✗ | ✓<br>*GBA* p.G241R P | ✗ | Prob. PSP-RS<br>O1, P0, A3, C0 | ✓ | ✓ |
| 11. Patient #4 | ✗ | ✓<br>*GBA* p.R89Q LP | ✗ | Prob. PSP-RS<br>O1, P1, A2, C0 | ✗ | ✗ |
| 12. Patient #37 | ✗ | ✓<br>*SYNJ1* p.A512V P | ✗ | Prob. PSP-RS<br>O1, P1, A2, C0 | ✗ | ✗ |
| 13. Patient #63 | ✗ | ✓<br>*SYNJ1* p.R284K LP | ✗ | Prob. PSP-RS<br>O1, P1, A3, C0 | ✗ | ✗ |
| Excluded Patients |  |  |  |  |  |  |
| Patient #6 | ✓<br>Moderate-to-severe cerebral atrophy and moderately severe leukoencephalopathy (note: had history of HTN) | *NOTCH3* p.R544C (the commonest variant associated with CADASIL in Asian patients including Chinese) | ✗ | Poss. PSP-RS<br>O1, P2, A2, C0 | ✗ | ✗ |
| Patient #96 | ✓<br>Confluent cerebral deep white matter T2 hyperintensity, with numerous microbleeds, raising<br>the possibility of<br>CAA | N/A | ✗ | Prob. PSP-RS<br>O1, P2, A2, C0 | ✗ | ✗ |

\*All mutations listed here were heterozygous and confirmed by Sanger sequencing. \*\*As per MDS-PSP clinical diagnostic criteria publication (OPAC=Oculomotor dysfunction, Postural Instability, Akinesia, Cognitive dysfunction). CAA=Cerebral amyloid angiopathy; CADASIL=Cerebral autosomal dominant arteriopathy with subcortical infarcts and leukoencephalopathy; F=Female; HTN=Hypertension; M=Male; MND=Motor neuron disease; N/A=Not available; LP=Likely pathogenic according to American College of Medical Genetics and Genomics (ACMG) criteria; P=Pathogenic; PD=Parkinson's disease; Poss.=Possible; Prob.=Probable; S.o.=Suggestive of; VP=Ventriculo-peritoneal; ✖=Negative/Absent; ✔=Positive/Present.

WES, performed in 48 (46.2%) patients, revealed five patients carrying heterozygous mutations in genes implicated in neurodegenerative disease (*GBA* in 3, and *SYNJ1* in 2), classified by American College of Medical Genetics and Genomics (ACMG) criteria as “pathogenic” or “likely pathogenic” (Table 3). These patients had motor symptom onset ranging 54-73 years; none had a family history of neurodegenerative disorders. No other pathogenic mutations in genes associated with “atypical PSP” presentations [3,7,8] were detected in the other (n=43) patients tested. One patient excluded on the basis of leukoencephalopathy on brain MRI was found to have a p.R544C mutation in *NOTCH3*, in keeping with a diagnosis of cerebral autosomal dominant arteriopathy with subcortical infarcts and leukoencephalopathy (CADASIL), which rarely has been shown to produce a PSP phenotype [3]. Further details of genetic findings will be reported in a separate publication.

A re-analysis done excluding all the patients with atypical imaging findings and/or pathogenic/likely pathogenic mutations did not materially change any of the main outcomes presented in Tables 1 and 2, except that the correlation between the PSP-CDS and LEDD became insignificant (P=0.056).

## 4. Discussion

In this study, we were able to classify all 104 consecutively recruited patients using the 2017 MDS-PSP clinical diagnostic criteria [3]. Our multi-ethnic cohort comprising Chinese, Indians and Malays showed a broadly similar spectrum of PSP predominance-types to that generally seen in European movement disorder clinics [2,9-11], with PSP-RS being the most common (accounting for ≈½ the cases), followed by PSP-P in ≈one-third and then PSP-PGF. However, since our study was performed in clinics with a focus on movement disorders, non-movement disorder presentations of PSP were likely under-represented (presenting instead to cognitive disorder clinics for example). Indeed, we did not diagnose any frontal (PSP-F) cases (which are not uncommon in some cohorts [12]), and encountered only one patient with PSP-SL [3].

Our cases underwent careful clinico-radiographic assessments by an experienced movement disorders neurologist, and next-generation (whole-exome) sequencing was done in ≈½ the cohort. These comprehensive evaluations resulted in the exclusion [3] of two patients with clinical PSP syndromes, one who probably had CADASIL and another who likely also had a vascular aetiology for her PSP syndrome. Nevertheless, it is known that while a diagnosis of PSP-RS is typically highly predictive of PSP pathology, other phenotypes such as PSP-P, PSP-PGF or PSP-CBS do not carry the same degree of diagnostic certainty and not infrequently are associated with other pathologies [1].

Interestingly, 10% of patients who had WES done in our cohort (all five were classified as probable PSP-RS [3]) carried gene variants associated with PD. However, none showed PD-suggestive features in the motor or non-motor domains (e.g., typical parkinsonian resting tremor, dramatic beneficial response to dopaminergic therapy, or levodopa-induced dyskinesias) [13], except for one patient harbouring a pathogenic *GBA* mutation (who had resting tremors, a history of RBD symptoms and visual hallucinations, as well as a supranuclear downgaze palsy - see Supplementary Video, Patient #18). Although mutations in *GBA* and *SYNJ1* are listed as exclusionary in the MDS-PSP diagnostic scheme (due to their links with PD, as well as DLB in the case of *GBA* [3]), there is emerging research implicating *GBA* variants in PSP patients [14,15]. This is perhaps unsurprising, given their role in lysosomal dysfunction involved in various neurodegenerative disorders. It is noteworthy, in this regard, that while variants in *LRRK2* were initially described to cause monogenic PD (as well as modulating the risk for sporadic PD), subsequent research linked some of the same mutations (p.G2019S and p.R1441C) with PSP-like phenotypes and pathology [16]. The findings of a recent large study involving ≈1,000 PSP patients of European ancestry (a large proportion of whom were neuropathologically confirmed) further support an important role for *LRRK2* in this condition [17]. There are also rare reports of homozygous mutations in *GBA* and *SYNJ1* causing familial atypical PSP syndromes (e.g., with onset in early adulthood) [8], but our patients with heterozygous variants were dissimilar: all had older age at onset (mean 64.6 years, similar to sporadic PSP) and lacked familial history of neurodegenerative disease.

Likewise, the unusual or unexpected neuroimaging findings in an additional eight patients were not considered sufficient to negate the diagnosis of PSP [3] and inclusion into our study. In any case, our main findings were unchanged when patients with the above genetic and neuroimaging findings were excluded from the analyses.

To date, there is only one PSP-specific rating scale that is “recommended” by the MDS for the global assessment of PSP severity [18]. The PSP Rating Scale (PSPRS), which consists of 28 items, is however time-consuming and may not be easily administered within the constraints of a busy clinic practice [19]. We found that the PSP-CDS could be conveniently administered as part of routine clinical care. The PSP-CDS scores in our cohort were comparable to that reported in the original publication (median 10.0 [IQR 7.0]; range 3-21; disease duration 4.7 [IQR 4.3] years in our study; vs. mean 10.8±3.6; range 5-20; disease duration 4.4±2.1 years).

In our cohort, PSP-CDS scores worsened with increasing disease duration, as expected for a progressive neurodegenerative disorder. A more modest, but significant, correlation was also seen between PSP-CDS and ZBI scores. This relatively low magnitude of effect was anticipated, given that many variables other than disease severity influence caregivers’ perception of burden (depending on psychological coping mechanisms, support from family members or paid help, etc.). Also, although seemingly paradoxical, a more advanced disease stage (with higher PSP-CDS scores) can sometimes be associated with *reduced* burden to caregivers, at least for certain aspects like falls, as a patient becomes non-ambulant. The elements we report here supporting construct validity of the PSP-CDS build on the high correlations found by the original authors between the PSP-CDS and other physician- and patient-reported outcomes [4]. We believe that the continued development and application of such tools as the PSP-CDS will be invaluable in helping to close the very wide gap in phenotypic characterization of movement disorders, especially in under-resourced settings [19].

We were able to find only two other publications that evaluated caregiver burden in PSP using the ZBI [20,21]. The larger study from India (n=77) reported a mean ZBI score of 21.9±12.5 [20]. This study involved patients with a relatively shorter duration of illness (mean 2.7±2.1 years) than ours [20]. Our higher ZBI scores (33.0 ± 15.7) thus were likely partly explained by longer duration (4.7 [IQR 4.3] years) of disease. The other study from Germany (n=20) involved patients with an even longer disease duration (6.3±3.0 years) and reported correspondingly higher ZBI scores (42.8±8.7) [21]. The overall high ZBI scores in PSP are in line with the contention that the levels of symptoms and caregiver burden in Parkinson-plus syndromes and advanced PD may be similar to - or even exceed - those of people entering palliative care with cancer [22], and underscore the need for urgent measures to be undertaken to improve supportive care for patients and families living with these conditions [23].

Some comment is warranted regarding several other clinico-demographic findings in our study. First, a relatively high rate (23.5%) of RBD symptoms was noted. Acknowledging that our assessment lacked polysomnographic confirmation, we took a conservative approach in endorsing the presence of RBD symptoms only when it was clinically convincing (e.g., sleep-talking alone did not qualify, in keeping with the RBD Single Question Screen [RBD1Q] and the Mayo Sleep Questionnaire [MSQ] core question regarding dream enactment) [24]. However, this still would not exclude the possibility that some cases could have been mimicked by non-REM sleep parasomnias, arousals from other sleep disorders such as sleep apnoea, or nocturnal confusion [24]. On the other hand, RBD is usually diagnosed and managed clinically in most practice settings. It is also now beginning to be recognized that RBD occurs not only in PD, DLB and MSA, but has also been polysomnographically-confirmed in cases of pathologically-proven non-synucleinopathy disorders, including PSP [25].

Second, we similarly found the prevalence of visual hallucinations (22.8% overall, and as high as 28.9% in PSP-P) to be substantially greater than that typically quoted for PSP (7% in the seminal study by Williams et al.) [26]. Visual hallucinations that are “predominant” are an exclusion criterion in the MDS-PSP diagnostic scheme [3], but this was not the case in our hallucinating patients, of whom only a minority (21.7%) required treatment with low-dose quetiapine (median dosage of 25mg/d). The genesis of visual hallucinations is believed to involve both disease and medication-related factors, with disease usually playing a greater role, at least in PD [27]. In our study, almost 80% of patients with a history of visual hallucinations were taking neither amantadine nor dopamine agonist. Notably, among the patients undergoing genetic testing in our cohort, no pathogenic mutation in genes such as *DCTN1, PGRN, C9orf72*, and *ATP13A2*, that can cause atypical parkinsonian disorders associated with hallucinations [7,8], was detected.

An association has been documented between RBD and hallucinations in PD as well as narcolepsy [24], and a moderate strength of correlation was likewise observed in our study. The occurrence of RBD and visual hallucinations (features typically associated with synucleinopathies) in our PSP patients could perhaps be explained, in part, by overlapping neuropathologies, which are common in older subjects [1]. For example, in a study evaluating 64 PSP cases diagnosed clinico-pathologically, 20% had concomitant PD pathology (conversely, concomitant PSP pathology was observed in 9% of 140 PD cases) [28]. For this, and other reasons (Supplementary Figure 1), conceptually it seems reasonable to expect some overlap in clinical features between PSP and PD.

Third, there has long been a suggestion in the literature that atypical parkinsonian disorders with poor levodopa responsiveness may be more common in people of Indian ancestry [29,30]. Whether this might relate to higher rates of Parkinson-plus syndromes such as PSP, or greater axial motor impairment because of a higher burden of cardiovascular risk factors [29], has not been adequately studied. In this context, it was interesting to note that the proportion of Indians in our PSP cohort (accounting for 21.2% of patients overall, and 26.0% of PSP-RS) seemed higher than that reported for consecutively-sampled PD patients in the same clinics (13.6% of 828 patients) [19]. Ethno-geographic factors likely contribute significantly to the heterogeneity of movement disorder presentations, but have not been adequately studied [30].

Finally, for what is usually regarded as a “sporadic” disorder [1], a possible family history of neurological conditions was relatively commonly reported, in 20.4% of our cohort. That said, we did not find any families with >1 person afflicted with PSP. Another caveat is that we did not systematically examine these family members, although interestingly in some instances we have personally evaluated and managed family members of PSP patients who fulfilled clinical diagnostic criteria for PD [13] (unpublished data).

Our study has several limitations. First, the study lacked pathological confirmation, which remains the gold standard for PSP diagnosis. Second, as with other cross-sectional studies, it is important to consider survivor bias, in which there is an over-representation of those who live longer (e.g., several of our PSP-P and PSP-PGF patients were still alive ≥10 years into their disease, ranging up to 17 years). This was reflected in a significantly shorter disease duration for PSP-RS (median 4.0 years vs. PSP-P [6.6 years] and PSP-PGF [6.2 years]), who are known to have a more aggressive disease course and poorer survival [1,2]. Therefore, the 48.1% period prevalence of PSP-RS may underestimate the true occurrence of these “classical” cases. Finally, we did not administer the more comprehensive PSPRS [18], as our objective was consecutive recruitment during routine clinic visits. This helped to avoid sampling bias, since severely disabled individuals are often excluded from research because of their inability to complete more laborious evaluations [19]. We note that Piot and colleagues previously demonstrated a high correlation between the PSP-CDS and the PSPRS (r=0.88, p<0.0001) [4].

In conclusion, we report on a well characterized prospective multi-ethnic Asian PSP cohort using the MDS-PSP diagnostic criteria and PSP-CDS rating scale. We observed higher-than-expected frequencies of RBD, visual hallucinations and positive family history, suggesting differential ethnic and possibly genetic susceptibilities to PSP and these complications, and important considerations when assessing Asian patients. We identified *GBA* and *SYNJ1* pathogenic variants in our cohort suggesting that variants in PD-linked genes can present with PSP features across different races.

## Data Availability

All data produced in the present study are available upon reasonable request to the authors.

## Acknowledgements

The authors gratefully acknowledge the generous participation of patients and their caregivers at the University of Malaya.

## Disclosures

### Ethical Compliance Statement

We confirm that we have read the Journal’s position on issues involved in ethical publication and affirm that this work is consistent with those guidelines.

### Financial Disclosures for the Previous 12 months

SYL receives grants from the Michael J. Fox Foundation. He serves on the Editorial Board for the Lundbeck Institute Neuroscience Foundation. He has received honoraria for talks sponsored by Medtronic, the International Parkinson & Movement Disorder Society (MDS), and the Shanghai Ruijin Forum for

### Translational Neurodegeneration

AHT receives grants from the Michael J. Fox Foundation, Ministry of Education Malaysia Fundamental Research Grant Scheme, and Toray Science Foundation Science & Technology Grant. She has received honoraria for talks sponsored by Sanofi, and the International Parkinson & Movement Disorder Society (MDS). PMM has received honoraria from National School of Public Health (ISCIII), Editorial Viguera and Takeda Pharmaceuticals for lecturing in courses; and from the International Parkinson and Movement Disorder Society (MDS) for management of the Program on Rating Scales.

---

**Supplementary Fig. 1.**
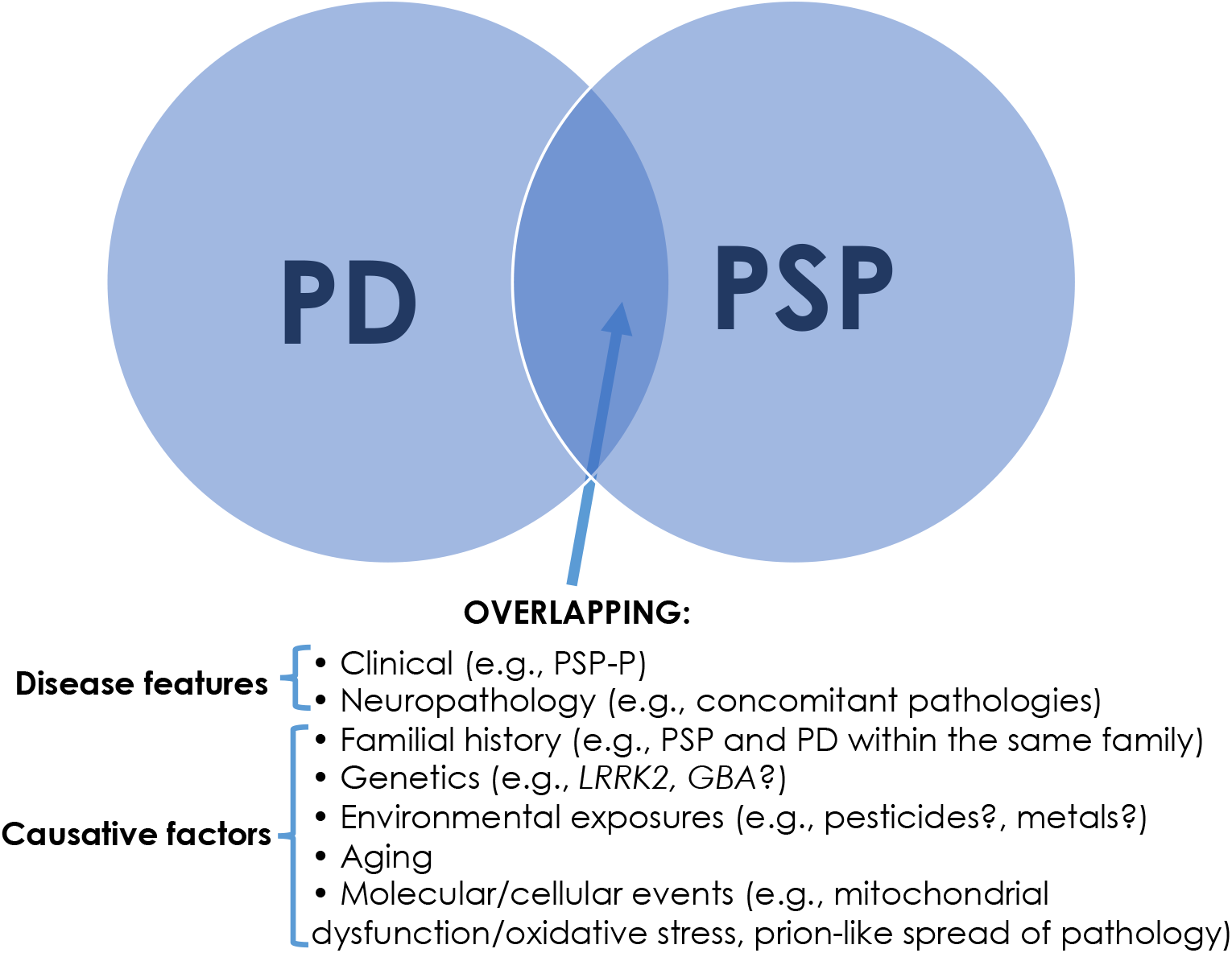
Areas of overlap between Parkinson’s disease (PD) and progressive supranuclear palsy (PSP).

